# Tracking and Predicting COVID-19 Epidemic in China Mainland

**DOI:** 10.1101/2020.02.17.20024257

**Authors:** Haoxuan Sun, Yumou Qiu, Han Yan, Yaxuan Huang, Yuru Zhu, Song Xi Chen

## Abstract

By proposing a varying coefficient Susceptible-Infected-Removal model (vSIR), we track the epidemic of COVID-19 in 30 provinces in China and 15 cities in Hubei province, the epicenter of the outbreak. It is found that the spread of COVID-19 has been significantly slowing down within the two weeks from January 27 to February 10th with 87.0% and 84.3% reductions in the reproduction number *R*_0_ among the 30 provinces and 15 Hubei cities, respectively. This suggests the extreme control measures implemented since January 23, which include cutting off Wuhan and many other cities and towns, a great public awareness and high level of self isolation at home, have contributed to a substantial decline in the reproductivity of the COVID-19 in China. We predict that Hubei province will reach its peak between February 20 and 22, 2020, and if the removal rate can be increased to 0.1, the epidemic outside Hubei province will end in May 2020, and inside Hubei in early June.

## 1. Introduction

The Corona Virus Disease 2019 (COVID-19) has created a profound public health emergency in China and has spread to 25 countries so far [1]. It has become an epidemic with more than 71,000 confirmed infections and 1,775 reported deaths worldwide as on February 17 2020. The COVID-19 is caused by a new corona viruses that is genetically similar to the viruses causing severe acute respiratory syndrome (SARS) and Middle East respiratory syndrome (MERS). Despite a relatively lower fatality rate comparing to SARS and MERS, the COVID-19 spreads faster and infects much more people than the SARS-03 outbreak.

The city of Wuhan, the origin of the outbreak, has been locked up to curtail population movement since January 23 in an effort to stop the spread of the epidemic, followed by more than 50 prefecture level cities (as on 8th of February) and countless number of towns and villages in China. A high percentage of the population are exercising self-isolation in their homes. The spring festival holiday period had been extended with all schools and universities closed and all students staying where they are indefinitely. The country is virtually in a stand-still, and the economy and people’s livelihood have been severely affected by the epidemic.

There is an urgent need to assess the speed of the disease transmission and to check if the existing containment measures have successfully slowed down the spread of the disease or not. The Susceptible-Infected-Removal (SIR) model [2] and its generalizations, for instance the SEIR model [3] with four or more compartments are commonly used to model the dynamics of infectious disease outbreaks. See [4, 5, 6, 7] for statistical estimation and inference for stochastic versions of the SIR model. SEIR models have been used to produce early results on COVID-19 in [8, 9, 10], which produced the first three estimates of the all important basic reproduction number *R*_0_: 2.68 by [8], 3.81 by [9] and 6.47 by [10]. The *R*_0_ is the expected number of infections by one infectious person during the course of his/her infectious period, and is a key measure of an epidemic. If *R*_0_ < 1, the epidemic will die down eventually with the speed of the decline depends on how small *R*_0_ is; otherwise, the epidemic will explode until it runs out of its course.

The SEIR models that was employed in the above three cited works for the COVID-19 assume constant model coefficients, implying a constant regime of transmission during the course of the epidemic. This is too idealistic for modeling COVID-19 as it cannot reflect the intervention measures by the authorities and the citizens, which should have made the infectious rate (*β*) and the reproduction number (*R*_0_) varying with respect to the time.

To reflect the changes due to the strong government intervention and self protection, we propose a varying coefficient SIR (vSIR) model, which can capture the varying dynamics of the epidemic. The vSIR model is easy to be implemented via the locally weighted regression approach [11] that produces estimates with desired smoothness, and yet is able to capture the changing dynamics of COVID-19’s reproduction, with guaranteed statistical consistency and needed standard errors. The consistent estimator and its confidence interval are needed for estimating the trend of *R*_0_, assessing the effectiveness of infection control (*R*_0_ is significantly less than 1 continuously for 7 days), and predicting the final number of infection cases and the future epidemic trend.

## 2. Results

By applying the vSIR model, we produce daily estimates of the infectious rate *β*(*t*) and the reproduction number *R*_0_(*t*) (*t* denotes time) for 30 provinces and 15 major cities (including Wuhan) in Hubei province from January 21 or a later date between January 24-29 depending on the first confirmed case to February 10. We report standard error in the parentheses following the estimate.

### 2.1. Main findings

- Despite the total number of confirmed cases and the death are increasing, the spread of COVID-19 has shown a great slowing down in China within the two weeks from January 27 to February 10 as shown by 88.0% and 86.8% reductions in the reproduction number *R*_0_ among the 30 provinces and the 15 cities in Hubei, respectively.
- The average 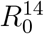 (based on 14-day infectious duration) on January 27th was 6.42 (1.57) and 7.67 (2.46), respectively, for the 27 provinces and the 7 Hubei cities with confirmed cases by January 23rd. One week later on February 3rd, the 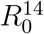 was averaged at 2.39 (0.70) for the 30 provinces and 2.94 (0.56) for the 15 Hubei cities, representing 62.8% and 61.7% reductions, respectively, over the 7 days. On February 10th, the average 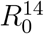 dropped further to 0.77 (0.33) for the 30 provinces and 1.01 (0.43) for the 15 Hubei cities, which were either below or close to the critical threshold level 1.
- The profound slowing down in the reproductivity of COVID-19 can be attributed to a series of action and measures by the government and the public, which include cutting off Wuhan and other cities from January 23, a rapid public awareness of the epidemic and the extensive self protection taken and high level of self isolation at home exercised over a much extended Spring Festival holiday period.
- There are increasing numbers of provinces and cities in Hubei whose 14-day *R*_0_ has been statistically below 1, as detailed in Table 1, which would foreshadow the coming of the turning point for containment of the epidemic, if the control measures implemented since January 23 can be continued.
- If the current decreasing trend of *R*_0_ continues, Hubei will reach peak infection between February 20 and February 22. Many non-Hubei provinces have already reached the peak. If the recovery rate can be increased to 0.1 meaning the average recover time is 10 days after diagnosis, the number of infected patients *I*(*t*) will be dramatically reduced in March, and the epidemic will end in early June; see Figure 3.
- The eventual control of COVID-19 is rested on if the existing control measures can be continued further for a period of time. The biggest challenges that can jeopardize the great effort from late January are from the impatient populations eager to get out of the self-isolation driven by either economic needs (migrant workers eager to coming back to cities for income) or people trying to escape from the boredness of self isolation while encouraged by the declining infections in the last two weeks.
- The implications of China’s experience in combating COVID-19 to other countries facing the epidemic are two folds. One is to reduce the person-to-person contact rate by self isolation and curtailing of population movement; another is to reduce the transmission probability by wearing protective wears should a contact has to be made.

**Table 1:** The reproduction number 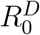 at three infectious durations: *D* = 7, 10.5, 14, for the 30 mainland provinces and 15 cities in Hubei province on February 10th. The symbols + (−) indicate that the 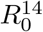 was significantly above (below) 1 at 5% level of statistical significance, and the numbers inside the square brackets were the consecutive days the 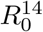 were above or below 1. The columns headed with Δ*R*_0_, Δ*R*_0_(1^st^) and Δ*R*_0_(2^nd^) are the percentages of decline in the 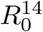 from the beginning of analysis to February 10th, to February 3rd, and the from February 3-10, respectively.

| Province/City | $R_0^7$ | $R_0^{10.5}$ | $R_0^{14}$ | $\Delta R_0$ | $\Delta R_0(1^{\text{st}})$ | $\Delta R_0(2^{\text{nd}})$ |
| --- | --- | --- | --- | --- | --- | --- |
| Ezhou | 0.9−[1] | 1.35+ | 1.8+ | 83.6% | 78.6% | 23.2% |
| Wuhan | 0.87−[1] | 1.31+ | 1.74+ | 71.2% | 43.7% | 48.8% |
| Tianmen | 0.87−[6] | 1.3+ | 1.74+ | 73.7% | 65.1% | 24.8% |
| Guizhou | 0.87−[4] | 1.3+ | 1.73+ | 62.8% | 8.3% | 59.4% |
| Hubei | 0.67−[2] | 1.01 | 1.34+ | 78.8% | 48.9% | 58.5% |
| Xiantao | 0.65−[2] | 0.97 | 1.3+ | 78.4% | 44.9% | 60.7% |
| Heilongjiang | 0.63−[2] | 0.95 | 1.26+ | 82.5% | 53.8% | 62.1% |
| Xinjiang | 0.6−[5] | 0.9−[2] | 1.2+ | 76% | 59.8% | 40.4% |
| Hebei | 0.58−[7] | 0.87−[1] | 1.16+ | 87.1% | 79.6% | 37% |
| Huangshi | 0.52−[3] | 0.78−[1] | 1.04 | 79.6% | 32.8% | 69.6% |
| Anhui | 0.47−[5] | 0.71−[1] | 0.95 | 89.1% | 70.9% | 62.6% |
| Shiyan | 0.46−[5] | 0.69−[1] | 0.91 | 89.7% | 71% | 64.4% |
| Jiangsu | 0.45−[5] | 0.68−[3] | 0.9 | 88.4% | 68.2% | 63.5% |
| Shandong | 0.45−[8] | 0.67−[2] | 0.9 | 91.3% | 82.9% | 49% |
| Yichang | 0.45−[6] | 0.67−[3] | 0.89 | 87.8% | 56.2% | 72% |
| Guangxi | 0.44−[8] | 0.66−[5] | 0.88 | 82.2% | 64.3% | 50.2% |
| Gansu | 0.43−[6] | 0.65−[1] | 0.87 | 82.2% | 48.6% | 65.5% |
| Shanxi | 0.43−[5] | 0.65−[3] | 0.86 | 88.9% | 66.6% | 66.8% |
| Tianjin | 0.43−[5] | 0.65−[2] | 0.86 | 84.5% | 52.6% | 67.3% |
| Xiangyang | 0.42−[5] | 0.63−[3] | 0.84−[1] | 88.6% | 57.3% | 73.3% |
| Jilin | 0.41−[3] | 0.62−[1] | 0.83 | 83.5% | 12.1% | 81.2% |
| Hainan | 0.41−[11] | 0.62−[2] | 0.82−[1] | 83.3% | 63.9% | 53.8% |
| Enshizhou | 0.41−[7] | 0.62−[4] | 0.82−[2] | 82.8% | 59.7% | 57.2% |
| Xiaogan | 0.39−[2] | 0.59−[1] | 0.79−[1] | 89.3% | 59.9% | 73.2% |
| Jingzhou | 0.38−[4] | 0.57−[2] | 0.76−[1] | 89.6% | 48.7% | 79.7% |
| Neimenggu | 0.38−[4] | 0.56−[3] | 0.75−[2] | 80.5% | 43.5% | 65.5% |
| Sichuan | 0.37−[7] | 0.55−[5] | 0.73−[3] | 91.1% | 77.9% | 59.9% |
| Jiangxi | 0.37−[4] | 0.55−[2] | 0.73−[1] | 91.9% | 69.2% | 73.8% |
| Suizhou | 0.35−[3] | 0.53−[2] | 0.7−[1] | 88.2% | 42.1% | 79.7% |

**Table 1 – continued from previous page**
| Province/City | $R_0^7$ | $R_0^{10.5}$ | $R_0^{14}$ | $\Delta R_0$ | $\Delta R_0(1^{\text{st}})$ | $\Delta R_0(2^{\text{nd}})$ |
| --- | --- | --- | --- | --- | --- | --- |
| Huanggang | 0.34–[4] | 0.52–[3] | 0.69–[1] | 91.6% | 58.4% | 79.7% |
| Jingmen | 0.34–[6] | 0.51–[1] | 0.68–[1] | 92.5% | 76.4% | 68.2% |
| Henan | 0.32–[4] | 0.48–[2] | 0.64–[1] | 94.4% | 77.3% | 75.1% |
| Beijing | 0.32–[6] | 0.47–[4] | 0.63–[1] | 89.7% | 60.9% | 73.8% |
| Hunan | 0.3–[5] | 0.46–[3] | 0.61–[2] | 93.6% | 74.7% | 74.9% |
| Shaanxi | 0.3–[7] | 0.45–[4] | 0.59–[3] | 88.9% | 60.3% | 72.1% |
| Chongqing | 0.29–[7] | 0.44–[4] | 0.58–[3] | 92.1% | 74.3% | 69.2% |
| Fujian | 0.27–[7] | 0.41–[5] | 0.55–[4] | 92.4% | 74.1% | 70.6% |
| Guangdong | 0.26–[5] | 0.39–[3] | 0.53–[2] | 90.2% | 51.4% | 79.8% |
| Liaoning | 0.26–[7] | 0.39–[6] | 0.51–[2] | 91.3% | 69.7% | 71.2% |
| Xianning | 0.22–[5] | 0.33–[3] | 0.45–[2] | 87.2% | 20.5% | 83.9% |
| Shanghai | 0.21–[7] | 0.32–[4] | 0.42–[3] | 92.6% | 64.2% | 79.3% |
| Ningxia | 0.2–[4] | 0.3–[2] | 0.4–[2] | 94.5% | 69.5% | 81.8% |
| Qinghai | 0.14–[5] | 0.21–[4] | 0.28–[4] | 89.8% | -1.4% | 89.9% |
| Yunnan | 0.14–[8] | 0.2–[7] | 0.27–[5] | 97.3% | 86.5% | 80.2% |
| Zhejiang | 0.14–[8] | 0.2–[4] | 0.27–[3] | 96.5% | 76.6% | 84.9% |
| Ave(sd) | 0.42(0.19) | 0.64(0.28) | 0.85(0.38) | 87.6% | 62.6% | 67% |

**Figure 1:**
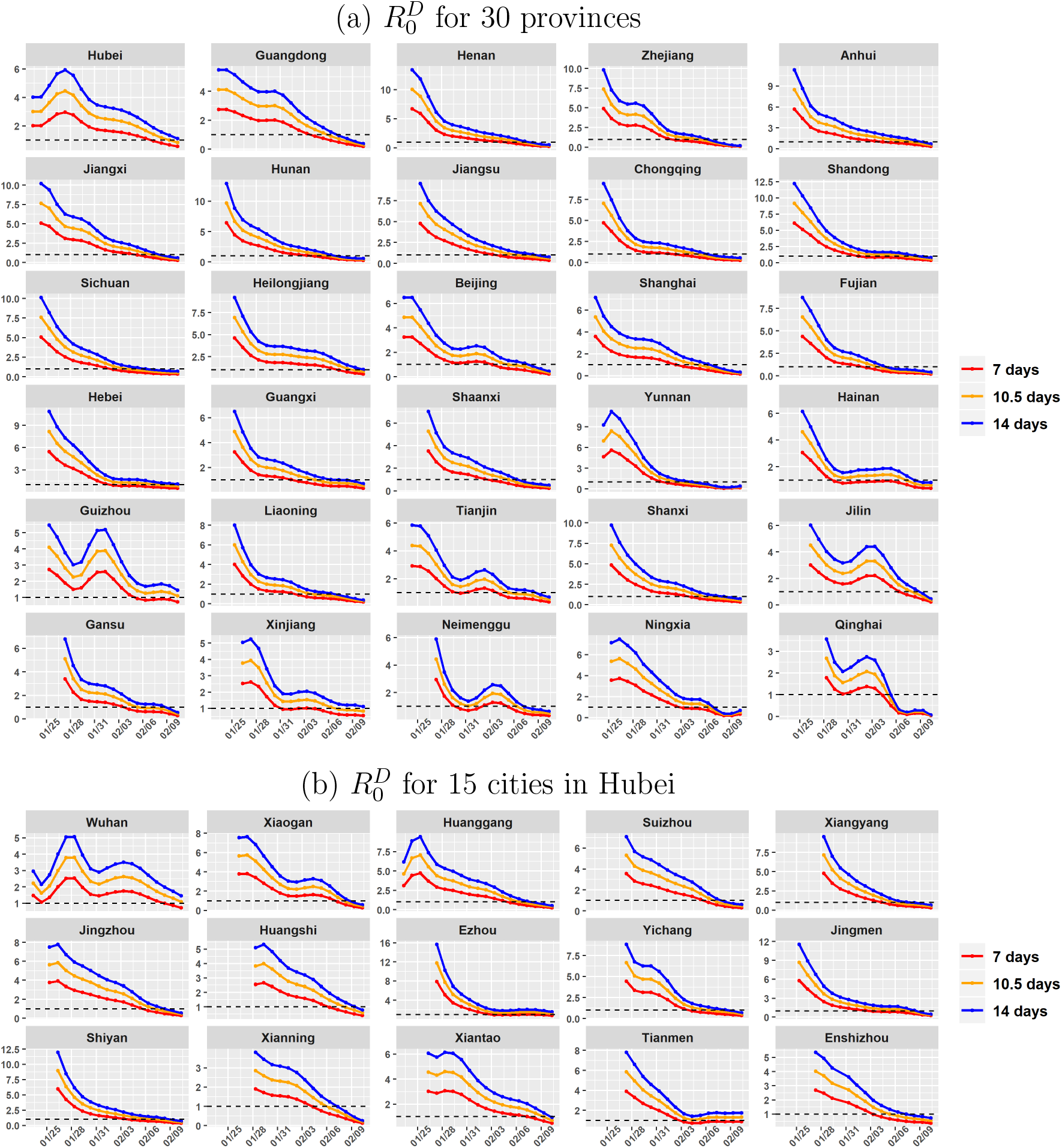
Time series of the reproduction number 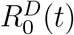 at three infectious durations: *D* = 7 (red), 10.5 (orange), 14 (blue), for the 30 mainland provinces (a) and the 15 cities in Hubei province (b) from Jan 21 to Feb 11 2020. The black horizontal line is the critical threshold level 1.

**Figure 2:**
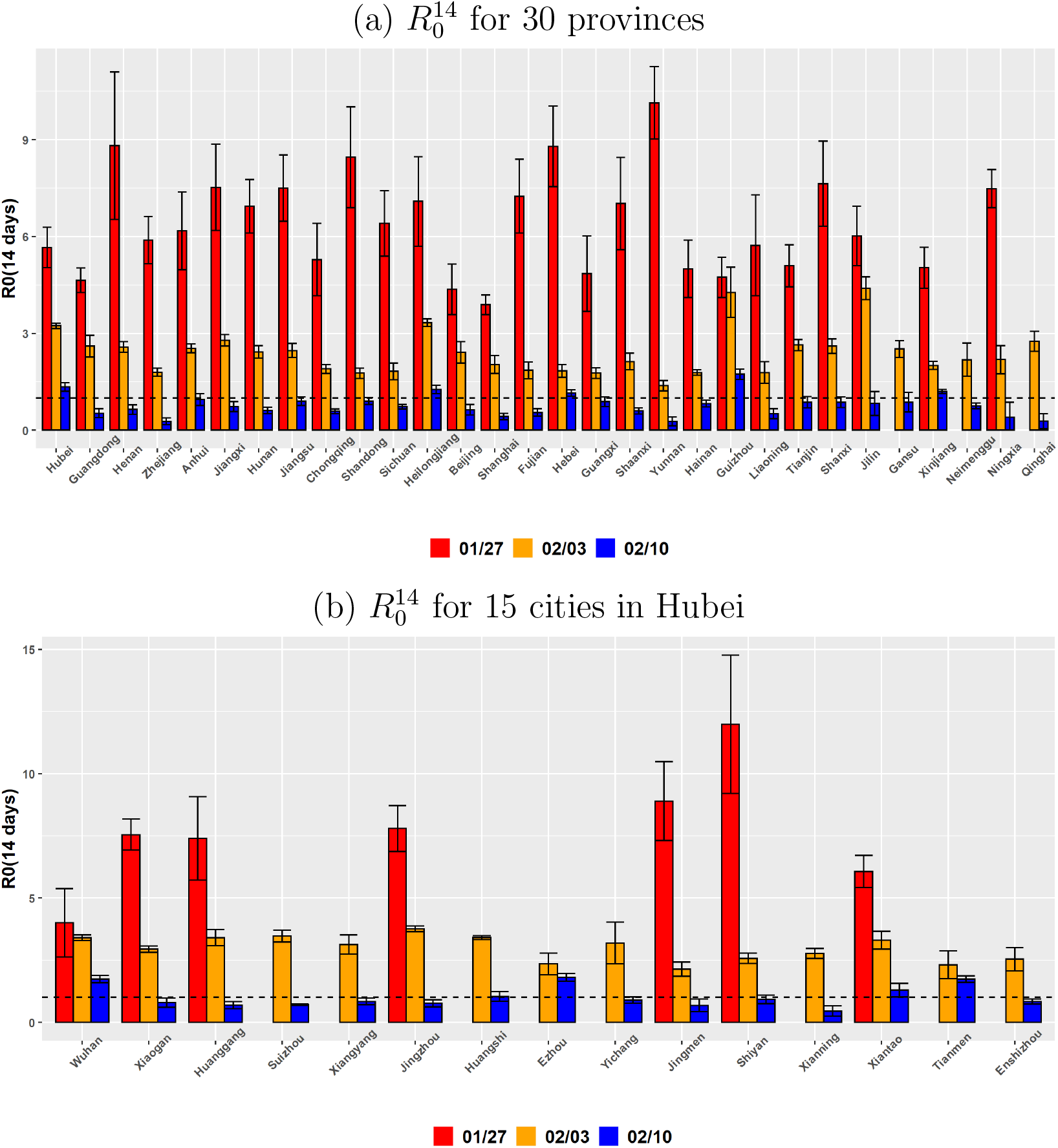
Elevated 95% confidence intervals (black) of the 14-day *R*_0_ for the 30 mainland provinces (a) and the 15 Hubei cities (b) on Jan 27 (red), Feb 3 (orange) and Feb 10 2020 (blue). The black horizontal lines mark the critical threshold 1.

**Figure 3:**
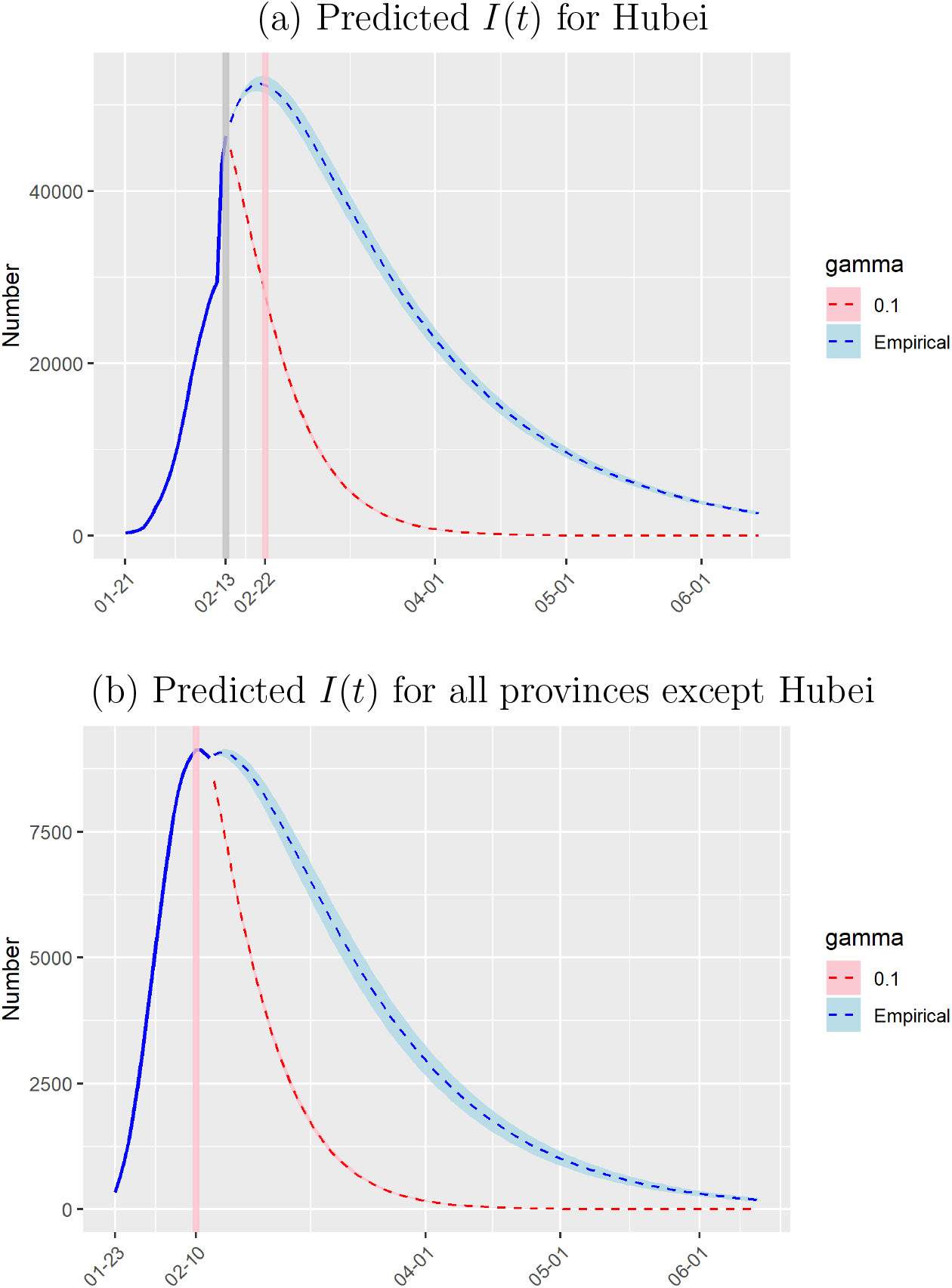
Predicted number of infected cases *I*(*t*) with 95% prediction interval for Hubei Province in panel (a) and all other provinces combined except Hubei in panel (b). The grey vertical line indicates the current date of observation; the blue solid line plots the observed *I*(*t*) before Feb 13th; the blue dashed line gives the predicted *I*(*t*) with 95% prediction interval (blue shaded area) with the estimated 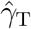 the pink vertical line indicates the peak date of *I*(*t*); the red dashed line gives the predicted *I*(*t*) with 95% prediction interval (red shaded area) with fixed recovery rate *γ* = 0.1.

### 2.2. Basic reproduction number

At a date *t*, the reproduction number based on an average infectious duration *D* is 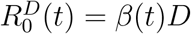 where *β*(*t*) is the daily infection rate at *t*. We do not adopt the version involving *γ*, the removal rate, since its estimation is highly volatile at the early stage of an epidemic. A general version of *R*0 (*t*) may be defined as 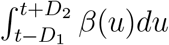 where positive *D* and *D* represent the infectious durations before and after diagnosis, respectively. The 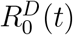 given above can be viewed as an approximation by the Middle Value Theorem in calculus with *D* = *D*_1_ + *D*_2_.

Research works [12, 13, 14] so far on COVID-19 have informed a range of duration for incubation, from onset of illness to diagnosis and then to hospitalization. The average incubation period from the three studies ranged from 3.0 to 5.2 days; the median duration from onset to diagnosis was 4 days [13]; and the mean duration from onset to first medical visit and then to hospitalization were 4.6 and 9.1 days [12], respectively. Based on a data sample of 391 cases from Shenzhen, the average incubation period was 4.46 (0.26) days and the average duration from onset to hospitalization were 3.9 (0.19) days, respectively, where standard error is reported in the parentheses. Another dataset of 100 confirmed cases in Shaoyang (Hunan Province) revealed the average durations from onset to diagnosis and from diagnosis to discharge were 5.67 (0.39) and 10.12 (0.43) days, respectively. There is a recent revelation [13] that asymptomatic patients can be infectious, which would certainly prolong the infectious duration.

There are much variation in the medical capability in timely diagnosis and hospitalization (thus quarantine) of the infected across the country. Thus, the infectious duration *D* would vary among the provinces and cities, and would change with respect to the stage of the epidemic as well.

Given the diverse range of infectious duration across the provinces and cities, in order to standardize and make the reproduction number *R*_0_ readily comparable, we calculated the 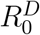 based on three levels of D: 7, 10.5 and 14 days, which represent three scenarios of responsiveness in diagnosing, hospitalization and hence quarantine of the infected. Calculation of the *R*_0_ at other duration can be made by inflating or deflating a 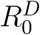 proportionally to reflect a local reality.

### 2.3. Reproductivity of COVID-19

Figures 1 presents the time series of 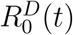 at the three levels of *D* for the 30+15 provinces/cities from late January to February 11th. Figure 2 displays three cross sectional 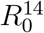 and their confidence intervals on January 27th, February 3rd and 10th, respectively.

Figure 1 reveals a monotone decreasing trend for almost all the provinces and cities with only exceptions for Hubei, Guizhou, Jinlin, Neimenggu and Qinghai. Even for those exceptional provinces, the recent trend is largely declining. The non-monotone pattern for non-Hubei provinces were largely due to relative small number of infected cases and waves of introduced infections. However, the one for Hubei and Wuhan suggests low data quality and in particularly under reporting and reporting delay. The epidemic statistics from Hubei and the city of Wuhan before January 21th were severely incomplete and with irregular patterns. This was the reason we start Hubei’s analysis from January 21th.

The average 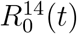 among the 27 provinces (with confirmed cases on and prior to January 23rd) was 6.42 (1.57), and 7.67 (2.46) for 7 of the 15 Hubei cities on January 27. These levels were comparable to the level of *R*_0_ (6.47) given in [10].

One week later on February 3rd, 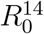 was averaged at 2.39 (0.70) for the 30 provinces and 2.94 (0.56) for the 15 Hubei cities, indicating that cutting off Wuhan and other cities, and the start of wearing face masks and self isolation at home from January 23th had contributed to 62.8% and 61.7% reduction in the *R*_0_. In the following week starting from February 4th, the average 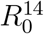 came down to 0.77 (0.33) for the 30 provinces and 1.01 (0.43) for the 15 Hubei cities on February 10th, representing further 67.8% and 65.6% reductions, respectively, during the second week. This reflects the beneficial effects of the continued large scale self-isolation within the extended spring festival holiday period.

Table 1 provides the reproduction number 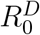 at the three durations on February 10th. It shows that 5 provinces and 4 Hubei cities’ 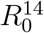 were significantly above 1 (at 5% significance level). There are 17 provinces and 8 Hubei cities’ 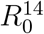 were significantly below 1, which were 4 and 6 more than those a day earlier on February 9th, and 9 and 8 more than those on February 8th, respectively. If we use the shorter *D* = 10.5, 27 provinces and 11 Hubei cities have been significantly below 1 for 1-7 consecutive days. These indicate that the reproduction number *R*_0_ has showed signs of crossing below the critical threshold 1 in increasing number of provinces and cities in Hubei around February 8-10. An updated Table 1 for February 16th are available in Table A1 in the Supplementary Information (SI), which shows continued improvement since February 10.

Given the significant decline in the reproduction numbers, it is time to discuss the turning point for COVID-19 for China. If a province or city’s 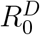 starts to be below 1 significantly (at 5% level), we would say the province or city have showed signs of the turning point. Given the uncertainty with the data records, especially those large variation in daily infected numbers coming out of Wuhan and Hubei, the turning point of the epidemic would be confirmed if 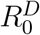 have been significantly below 1 for *D*_1_ days, where *D*_1_ is the period of infection before diagnosis, assuming all diagnosed can be quarantine immediately. Based on the results in [12, 13, 14], *D*_1_ = 7 may be considered. Then, some of the 30+15 provinces/cities have already reached the turning point, and more will be so in the coming days according to latest Table A1 in SI.

### 2.4. Prediction

Based on the estimated *β*(*t*) over time, we predict COVID-19’s future trajectories as solutions to the vSIR model. We consider two scenarios for the recovery rate *γ*. One uses the empirical estimate based on data to February 13th. As an effective cure for the virus has not been found, the estimated recovery rates are quite low. Among the provinces with more than 100 infections on February 13, Hunan had the highest recovery rate 0.06, followed by Jilin and Zhejiang (0.05), and then Tianjin, Chongqing, Hebei, Guizhou, Henan and Shanghai (0.046–0.049). Hubei, the province at the center of the epidemic, was 0.021. The other scenario is to choose *γ* = 0.1, which means the average removal time from diagnosis is 10 days, representing improvement in the treatment for COVID-19 patients as time progress.

Tables 2 and 3 present the 95% prediction intervals for the peak and end times of the number of infections (subtracted by the number of removals), and the cumulative number of infected at the ending based on the two scenarios of the recovery rate, respectively. We use data to February 13 2020 for the prediction. The predicted infection number *Î*(*t*) is within 5% and 10% deviation from its observed value on February 14 and February 15, 16 respectively; see Table A2 in SI for the detailed prediction error. The prediction based on the most recent data to February 16 gives similar results.

**Table 2:**
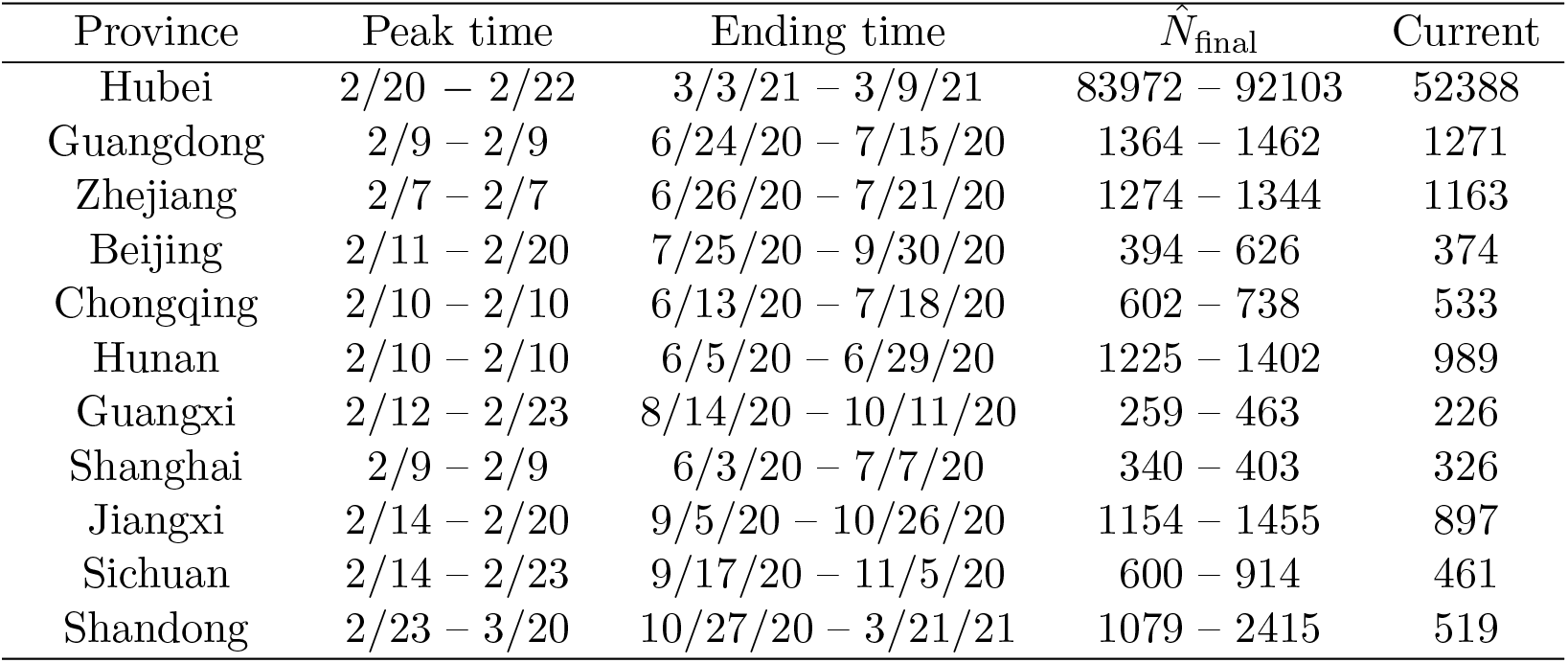
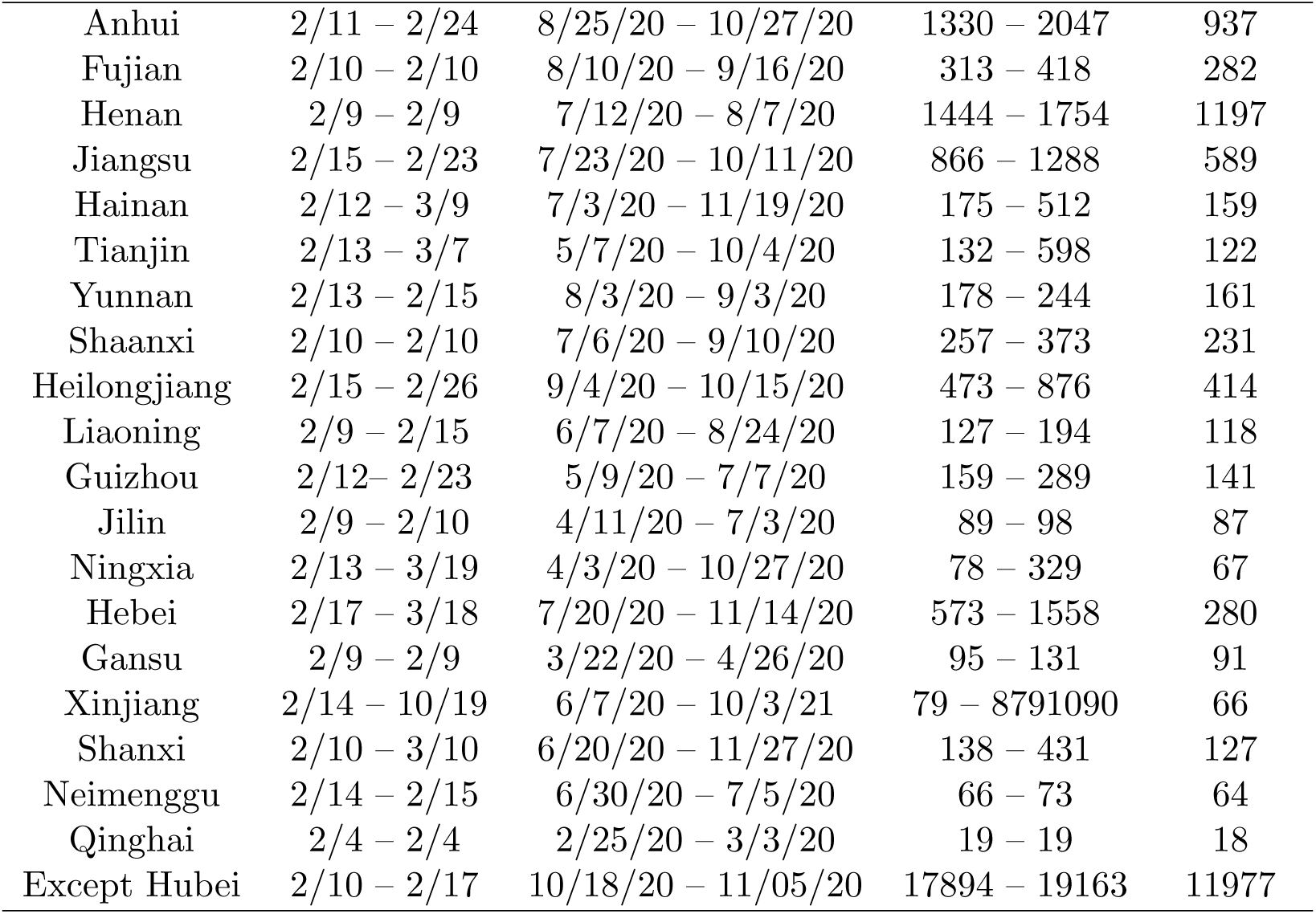
The 95% prediction intervals for the peak and ending times, and the final accumulative number of infected cases of COVID-19 epidemic in the 30 provinces based on data to Feb 13 2020 with the estimated 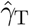. The last column lists the total infected cases (*I*(*t*) + *R*(*t*)) as Feb 13, 2020.

| Province | Peak time | Ending time | $\hat{N}_{\text{final}}$ | Current |
| --- | --- | --- | --- | --- |
| Hubei | 2/20 – 2/22 | 3/3/21 – 3/9/21 | 83972 – 92103 | 52388 |
| Guangdong | 2/9 – 2/9 | 6/24/20 – 7/15/20 | 1364 – 1462 | 1271 |
| Zhejiang | 2/7 – 2/7 | 6/26/20 – 7/21/20 | 1274 – 1344 | 1163 |
| Beijing | 2/11 – 2/20 | 7/25/20 – 9/30/20 | 394 – 626 | 374 |
| Chongqing | 2/10 – 2/10 | 6/13/20 – 7/18/20 | 602 – 738 | 533 |
| Hunan | 2/10 – 2/10 | 6/5/20 – 6/29/20 | 1225 – 1402 | 989 |
| Guangxi | 2/12 – 2/23 | 8/14/20 – 10/11/20 | 259 – 463 | 226 |
| Shanghai | 2/9 – 2/9 | 6/3/20 – 7/7/20 | 340 – 403 | 326 |
| Jiangxi | 2/14 – 2/20 | 9/5/20 – 10/26/20 | 1154 – 1455 | 897 |
| Sichuan | 2/14 – 2/23 | 9/17/20 – 11/5/20 | 600 – 914 | 461 |
| Shandong | 2/23 – 3/20 | 10/27/20 – 3/21/21 | 1079 – 2415 | 519 |
| Continued on next page |  |  |  |  |

**Table 2 – continued from previous page**
| Province | Peak time | Ending time | $\hat{N}_{\text{final}}$ | Current |
| --- | --- | --- | --- | --- |
| Anhui | 2/11 – 2/24 | 8/25/20 – 10/27/20 | 1330 – 2047 | 937 |
| Fujian | 2/10 – 2/10 | 8/10/20 – 9/16/20 | 313 – 418 | 282 |
| Henan | 2/9 – 2/9 | 7/12/20 – 8/7/20 | 1444 – 1754 | 1197 |
| Jiangsu | 2/15 – 2/23 | 7/23/20 – 10/11/20 | 866 – 1288 | 589 |
| Hainan | 2/12 – 3/9 | 7/3/20 – 11/19/20 | 175 – 512 | 159 |
| Tianjin | 2/13 – 3/7 | 5/7/20 – 10/4/20 | 132 – 598 | 122 |
| Yunnan | 2/13 – 2/15 | 8/3/20 – 9/3/20 | 178 – 244 | 161 |
| Shaanxi | 2/10 – 2/10 | 7/6/20 – 9/10/20 | 257 – 373 | 231 |
| Heilongjiang | 2/15 – 2/26 | 9/4/20 – 10/15/20 | 473 – 876 | 414 |
| Liaoning | 2/9 – 2/15 | 6/7/20 – 8/24/20 | 127 – 194 | 118 |
| Guizhou | 2/12 – 2/23 | 5/9/20 – 7/7/20 | 159 – 289 | 141 |
| Jilin | 2/9 – 2/10 | 4/11/20 – 7/3/20 | 89 – 98 | 87 |
| Ningxia | 2/13 – 3/19 | 4/3/20 – 10/27/20 | 78 – 329 | 67 |
| Hebei | 2/17 – 3/18 | 7/20/20 – 11/14/20 | 573 – 1558 | 280 |
| Gansu | 2/9 – 2/9 | 3/22/20 – 4/26/20 | 95 – 131 | 91 |
| Xinjiang | 2/14 – 10/19 | 6/7/20 – 10/3/21 | 79 – 8791090 | 66 |
| Shanxi | 2/10 – 3/10 | 6/20/20 – 11/27/20 | 138 – 431 | 127 |
| Neimenggu | 2/14 – 2/15 | 6/30/20 – 7/5/20 | 66 – 73 | 64 |
| Qinghai | 2/4 – 2/4 | 2/25/20 – 3/3/20 | 19 – 19 | 18 |
| Except Hubei | 2/10 – 2/17 | 10/18/20 – 11/05/20 | 17894 – 19163 | 11977 |

**Table 3:** The 95% prediction intervals for the peak and ending times, and the final accumulative number of infected cases of COVID-19 epidemic in the 30 provinces based on data to Feb 13 2020 with *γ* = 0.1. The last column lists the total infected cases (*I*(*t*) + *R*(*t*)) as Feb 13, 2020.

| Province | Peak time | Ending time | $\hat{N}_{\text{final}}$ | Current |
| --- | --- | --- | --- | --- |
| Hubei | 2/14 – 2/14 | 6/8 – 6/10 | 69896 – 73460 | 52388 |
| Guangdong | 2/9 – 2/9 | 4/24 – 4/26 | 1341 – 1413 | 1271 |
| Zhejiang | 2/7 – 2/7 | 4/23 – 4/25 | 1239 – 1286 | 1163 |
| Beijing | 2/11 – 2/11 | 4/12 – 4/25 | 390 – 513 | 374 |
| Chongqing | 2/10 – 2/10 | 4/16 – 4/24 | 577 – 675 | 533 |
| Hunan | 2/10 – 2/10 | 4/25 – 5/1 | 1151 – 1261 | 989 |
| Guangxi | 2/12 – 2/12 | 4/9 – 4/21 | 247 – 310 | 226 |
| Shanghai | 2/9 – 2/9 | 4/10 – 4/14 | 340 – 373 | 326 |

**Table 3 – continued from previous page**
| Province | Peak time | Ending time | $\hat{N}_{\text{final}}$ | Current |
| --- | --- | --- | --- | --- |
| Jiangxi | 2/13 – 2-13 | 4/25 – 4/29 | 1050 – 1153 | 897 |
| Sichuan | 2/13 – 2/13 | 4/18 – 4/26 | 537 – 617 | 461 |
| Shandong | 2/11 – 2/11 | 4/27 – 5/16 | 704 – 889 | 519 |
| Anhui | 2/11 – 2/11 | 4/28 – 5/6 | 1169 – 1360 | 937 |
| Fujian | 2/10 – 2/10 | 4/10 – 4/16 | 302 – 342 | 282 |
| Henan | 2/9 – 2/9 | 4/26 – 5/1 | 1362 – 1501 | 1197 |
| Jiangsu | 2/13 – 2/13 | 4/23 – 5/4 | 728 – 876 | 589 |
| Hainan | 2/12 – 2/12 | 4/4 – 4/15 | 172 – 225 | 159 |
| Tianjin | 2/12 – 2/12 | 4/2 – 5/11 | 134 – 257 | 122 |
| Yunnan | 2/13 – 2/13 | 4/5 – 4/10 | 172 – 192 | 161 |
| Shaanxi | 2/10 – 2/10 | 4/8 – 4/15 | 245 – 292 | 231 |
| Heilongjiang | 2/13 – 2/13 | 4/15 – 4/22 | 460 – 586 | 414 |
| Liaoning | 2/9 – 2/9 | 4/1 – 4/10 | 125 – 152 | 118 |
| Guizhou | 2/12 – 2/12 | 4/4 – 4/14 | 155 – 216 | 141 |
| Jilin | 2/9 – 2/9 | 3/27 – 3/29 | 89 – 95 | 87 |
| Ningxia | 2/13 – 2/13 | 3/29 – 6/2 | 78 – 166 | 67 |
| Hebei | 2/13 – 2/13 | 4/27 – 5/24 | 427 – 594 | 280 |
| Gansu | 2/9 – 2/9 | 3/26 – 4/10 | 94 – 124 | 91 |
| Xinjiang | 2/13 – 2/13 | 3/29 – 12/24 | 77 – 466 | 66 |
| Shanxi | 2/10 – 2/10 | 4/1 – 4/20 | 136 – 193 | 127 |
| Neimenggu | 2/13 – 2/13 | 3/26 – 3/28 | 65 – 73 | 64 |
| Qinghai | 2/4 – 2/4 | 3/3 – 3/7 | 19 – 19 | 18 |
| Except Hubei | 2/10 – 2/10 | 5/25 – 5/27 | 15158 – 15651 | 11977 |

From Table 2, with the estimated recovery rate, Hubei will reach peak infection between February 20 and February 22. For many non-Hubei provinces, their peak time have already occurred as early as February 4 (Qinghai), February 7 (Zhejiang) and February 9 (Guangdong, Shanghai, Henan, Jilin, Gansu), and five other provinces on February 10. The last row gives the predictions for all the non-Hubei provinces combined, which reaches peak infection between February 10th and 17th with 95% confidence.

From the trajectory of the vSIR model, the epidemic will end in late October 2020 for the non-Hubei provinces with the accumulated number of final infected cases in the range 17,894–19,163. The total non-Hubei infected number was 11,977 (as February 13). The ending time of Hubei is predicted to be March 2021 with final infected in the range 83,972–92,103. The current total infected cases in Hubei was 52,388 on February 13th.

It should be highlighted that the above prediction results were based on the estimated recovery rate so far. Table 3 gives the results with the recovery rate increased to 0.1. The trajectories of *I*(*t*) under the proposed vSIR model with the estimated recovery rate and *γ* = 0.1 are presented in Figure 3. With a higher recovery rate of 0.1, the duration of the epidemic will be shorten substantially. Figure 3 indicates that the number of infected will quickly decease in late February and March with very few cases left in April. The ending time for Hubei will be brought early to June 2020 with total number of infection reduced to the range 69,896–73,460, down by 14,076–18,643. Most of the non-Hubei provinces will end in April, 2020. Some provinces with few number of total infected cases may end as early as March (Qinghai, Jilin, Neimenggu). This shows that improving the recovery rate is an efficient way to end the COVID-19 infection early given the current decreasing trend of *β*(*t*), as it leads to the reduction of the infectious duration.

## 3. Methods

Let *S*(*t*), *I*(*t*) and *R*(*t*) be the counts of susceptible, infected and recovered (including dead) persons in a given city or province at time *t*, respectively. Let *N* be the total population of the city/province. We propose a varying coefficient Susceptible-Infected-Recovered (vSIR) model to estimate the dynamics of COVID-19 and predict its future course of spread.

### 3.1. Data

The daily records of infected, dead and recovered patients released by National Health Commission of China (NHCC) are obtained from the NHCC website, with the first confirmed record for Wuhan on December 8th, 2019, followed by 30 provinces in mainland China and 15 cities in Hubei province where Wuhan is the capital city. We did not consider data from Tibet due to very small number of cases. Table A3 in SI provides the starting dates of the data records and analysis for each province and city. Due to severe under-reporting in the first 39 days of the epidemics in Wuhan and Hubei, we consider data from January 16th for Wuhan and Hubei. For other provinces and Hubei cities, the starting dates for data are those of first confirmed case, and the analysis date started four days afterward due to the estimation approach for estimating the infectious rate *β*(*t*). The latest start for analysis was January 29th for Qinghai province and three cities in Hubei province. The second last date was January 28th that started 2 provinces and 5 Hubei cities.

The data from *Shenzhen Government Online* are epidemic statistics released by the Shenzhen Municipal Health Commission from January 19th to February 13th [15]. One dataset about the details of confirmed cases contains the time of onset, time of hospital admission, cause of illness and other information of 391 cases, including 188 males and 203 females. The admission time of these cases ranged from January 9th to February 11th. The other dataset reports the discharge time for 94 cases in the former dataset. Besides, the dataset of 100 confirmed cases was released by the Shaoyang Municipal Health Committee [16] on February 14 that includes 48 males and 52 females with the onset dates ranging from January 12 to February 11.

### 3.2. Time-varying coefficient SIR model

The Susceptible-Infective-Removal (SIR) model [2] is a commonly used epidemiology model for the dynamic of susceptible *S*(*t*), infected *I*(*t*) and recovered *R*(*t*) as a system of ordinary differential equations (ODEs). Here we consider a more generalized version of the SIR model in that the infectious rate *β* and the removal rate *γ* may change with respect to time so that

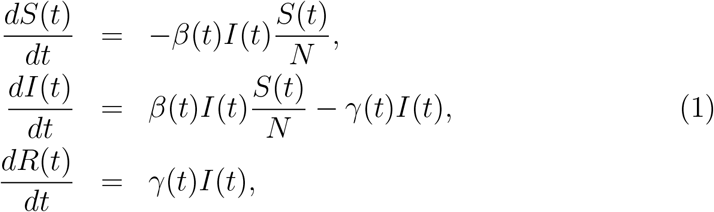

where *β*(*t*) and *γ*(*t*) are unknown functions of time.

The rationale for using a time-varying *β*(*t*) function, rather than a constant *β*, is that *β*(*t*) is the average rate of contact per unit time multiplied by the probability of disease transmission per contact between a susceptible and an infectious subject. Due to an increasing public awareness of the epidemic and the control measures put in place, both the transmission probability and the contact rate have been reduced due to protective wear (face mask), avoidance of close contacts and self isolation. These favors for a time-varying *β*(*t*) are also confirmed by the sharp declined in 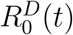 in Figures 1 and 2.

The removal rate will also change over time as treatments improve over time. However, our analysis (Figure S4 in SI) shows *γ*(*t*) is much slowly changing for most of the provinces, which led us to treat *γ*(*t*) = *γ* at current stage of the outbreak.

### 3.3. Estimation and inference

The reported numbers of infected and removed cases are subject to measurement errors. To reduce the errors, we apply a three point moving average filter on the reported counts to obtain *Ī*(*t*) = 0:3*I*(*t* − 1) + 0:4*I*(*t*)+0:3*I*(t+1) for 2 ≤ *t* ≤ *T* − 1 where *T* is the latest time point of observation. In our analysis, *T* is February 13 2020. For *t* = 1 or *T*, we apply two point averaging with 7*/*10 weight at *t* = 1 or *T*, and 3*/*10 for *t* = 2 or *T* − 1. Apply the same filtering on the recovered process *R*(*t*) and obtain 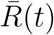. To simply the notation, we denote the filtered data *Ī*(*t*) and 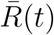 as *I*(*t*) and *R*(*t*) respectively, wherever there is no confusion.

Let Δ_*δ*_*R*_*t*_ = *R*_*t*+*δ*_ − *R*_*t*_ for *t* = 1, …, *T* − *δ*. From the third equation in (1), we estimate *γ* by least square fitting of Δ_*δ*_*R*_*t*_ on *I*(*t*) without intercept. We estimate *β*(*t*) − *γ* by a local linear regression on log{*I*(*t*)}. Let 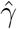 and 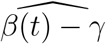 be the estimators, and 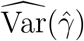 and 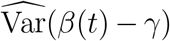 be their estimated variances. Their close form expressions are provided in Section S.1 in SI. Then, 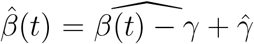 is the estimate for the varying coefficient *β*(*t*) in (1). The standard error of 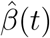 can be obtained as 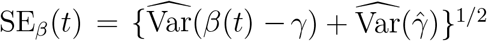. The 95% confidence interval for *β*(*t*) can be constructed as

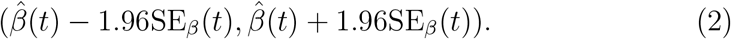

In the implementation, we chose *δ* = 2 and *w* = 5. Figure S1 in SI shows that the proposed vSIR model fits the observed infected number *I*(*t*) well for 30 provinces in China.

### 3.4. Prediction for infection rate and state variables

As 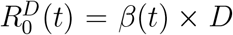, predicting *β*(*t*) is equivalent to predicting 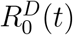. From Figure 1 and Figure S2 in SI, we see that the overall trends of *β*(*t*) is decreasing. But the rate of deceasing gets smaller as time travels. To model such trend, we consider the reciprocal regression

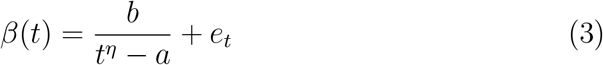

with error *e*_*t*_ and unknown parameters *a, b* and *η*. The parameters *a, b* and *η* are estimated by minimizing the sum-of-square distance between the estimates 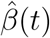 and their fitted values.Let 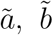 and 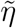 be the estimated parameters, and 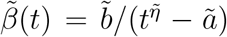 be the fitted function. Figure S3 in SI shows the reciprocal model fits 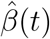 quite well for most of the provinces, especially those with large number of infected cases.

With the fitted 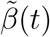, we project {*S*(*t*), *I*(*t*), *R*(*t*)} via the ODEs

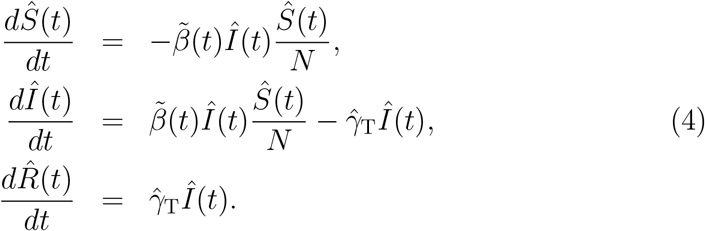

where 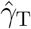 is the estimated recovery rate at time *T* using the last five days’ data. With the observed {*S*(*T*), *I*(*T*), *R*(*T*)} at the current time *T* as the initial values, numerical solutions 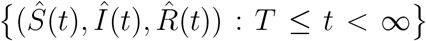 for the system (4) could be obtained using the Euler method. Then, the peak time of the number of infected cases can be predicted as *t*_peak_ = arg max*t Î*(*t*), and the estimated final infected number is 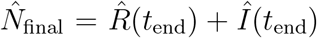, where *t*_end_ = min {*t* Î(*t*) < 1} is the estimated ending time. The 95% prediction intervals for the peak time, end time and final infected number are obtained by bootstrap resampling method. The details of the bootstrap prediction inference is provided in Section S.2 in SI.

## Data Availability

The data used for this study are publicly available. The daily records of infected, dead and recovered patients are released by National Health Commission of China (NHCC).

